# Psychological Legacies of Intergenerational Trauma under South African Apartheid: Prenatal Stress Predicts Increased Psychiatric Morbidity during Late Adolescence and Early Adulthood in Soweto, South Africa

**DOI:** 10.1101/2021.01.11.21249579

**Authors:** Andrew Wooyoung Kim, Rihlat Said Mohamed, Shane A. Norris, Linda M. Richter, Christopher W. Kuzawa

## Abstract

**Background:** South Africa’s rates of psychiatric morbidity are among the highest in sub-Saharan Africa and are foregrounded by the country’s long history of political violence during apartheid. Recent evidence suggests that maternal trauma during gestation may intergenerationally impact the developing fetus and elevate the future child’s risk for psychiatric disease. We aim to evaluate the intergenerational effects of prenatal stress experienced during apartheid on late adolescent psychiatric morbidity and also assess the potential ameliorative effects of prenatal social support.

**Method:** Participants (n = 1051) come from Birth-to-Twenty, a longitudinal birth cohort study in Soweto-Johannesburg, South Africa’s largest peri-urban township which was the epicenter of violent repression and resistance during the final years of the apartheid regime. Pregnant women were prospectively enrolled in 1990 and completed questionnaires assessing social experiences, and their children’s psychiatric morbidity were assessed at age 17-18.

**Results:** Full data were available from 304 mother-child pairs in 2007-8. Pregnant women who experienced worse traumatic stress in 1990 had children who exhibited greater psychiatric morbidity during late adolescence. This relationship was only significant, however, among children born to younger mothers and adolescents who experienced greater concurrent stress. Social support did not buffer the long-term impacts of prenatal stress on future psychiatric morbidity.

**Conclusion:** Greater prenatal stress predicted adverse psychiatric outcomes among children born to younger mothers and adolescents who experienced greater concurrent stress. Our findings suggest that prenatal stress may affect adolescent mental health, have stress-sensitizing effects, and represent possible intergenerational effects of trauma experienced under apartheid in this sample.

## INTRODUCTION

South Africa’s rates of mental, neurological, and substance use disorders are among the highest in sub-Saharan Africa. In 2016, the estimated 12-month prevalence for any psychiatric disorder was 16.2%, or approximately 9.1 million individuals (GBDCN 2017). Despite these elevated rates of psychiatric morbidity, access to mental health treatment is poor: only 27% of patients living with severe mental illness are expected to receive treatment (Herman et al. 2009). The current state of public mental health in South Africa is foregrounded by a long and recent history of institutionalized White supremacist policies implemented during the *Apartheid* regime (c. 1948-1994). This period was characterized by systematic disenfranchisement of non-White communities through various modes of social, economic, and political oppression (Beinhart 2001). Despite the legislative end of *Apartheid* policies and the birth of a new democratic in 1994, South African society continues to be plagued by the persistent societal institutions of *Apartheid*, including chronic poverty (Gibbs et al. 2018), discrimination (Kuzawa & Sweet 2009), and racialized class inequality (Adjaye-Gbewonyo et al. 2016; Burns 2015), all of which are known risk factors for psychiatric disorders (Allen et al. 2014; Moomal et a. 2009; Myer et al. 2008).

In addition to these ongoing societal effects of poverty, structural violence and inequality, there is growing evidence, from South African studies and other populations, that the stressors of the past could have lingering biological effects that continue to influence socio-emotional behavior and mental health across the lifecourse. Growing evidence from the fetal origins of health and disease framework shows that past stress and trauma exposures, particularly those that occur during early development, can durably alter the development and function of various stress regulatory mechanisms in humans, including the immune system, cardiovascular system, neurobiological function, and neuroendocrine pathways (Barker et al. 1999; Gluckman & Hanson 2004; Kuzawa 2008; Taylor et al. 2010; Heim et al. 2019). Recent findings also suggest that these stress-linked alterations in stress physiology may also affect the developing offspring through various pathways of genetic and non-genetic inheritance. For instance, large population-based studies in the United Kingdom show that greater levels of maternal prenatal stress are associated with an increased future risk for externalizing disorders in their children, such as attention deficit hyperactivity disorder (Rice et al. 2010), conduct disorders (MacKinnon et al. 2018), and internalizing disorders like anxiety and depression among adolescents (O’Donnell et al. 2014; Sharp et al. 2015). Additionally, the intergenerational signatures of maternal prenatal stress exposure have been reported in offspring during adulthood in large cohort studies and across various contexts such as Australia, the Philippines, and the United States (Betts et al. 2015; DeSantis et al. 2015; Entringer et al. 2009).

While the specific biological mechanisms that underlie the long-term psychiatric effects of prenatal stress are unknown, growing evidence from the literature on the fetal origins of psychopathology suggests that prenatal stress exposure alters the development, function, sensitivity of human stress physiological systems across the life course, which in turn may elevate one’s risk for a psychopathological presentation. These stress-sensitive systems include the immune system, the catecholamine-producing sympathetic-adrenal-medullary (SAM) axis, and the hypothalamic-pituitary-adrenal (HPA) axis, which regulates physiological reactions to stress through stress-sensitive hormones like cortisol. Maternal stress-induced elevations in cortisol can enter maternal circulation, penetrate the placental wall, and reach the gestational environment of future offspring, thereby impacting the development of fetal stress physiology (D’Anna-Hernandez et al. 2012; O’Donnell et al. 2009).

Greater fetal exposure to cortisol is understood influence a wide variety of biological and health phenotypes across the lifecourse. These include birth outcomes such as restricted fetal growth rates, shorten gestations, and reduce birth size (Kim et al. 2020a; Rosa et al. 2019; Ryu 2019). Greater intrauterine cortisol exposure may also durably alter the development and sensitivity of the fetal stress physiology (e.g. HPA axis, immune system, brain development), which together with the maternal and placental systems, may have sustained impacts on the child’s stress physiology across their lifecourse (Brand et al. 2010; DeSantis et al. 2015; Entringer et al. 2009; Karlén et al. 2013). Early life stress-linked dysregulation of stress physiological mechanisms, characterized by altered diurnal cortisol rhythms, glucocorticoid resistance, chronic low-grade inflammation, and modulated brain function, has consistently been reported as both a prospective risk factor and cross-sectional neuropsychiatric phenotype of a range of psychiatric illnesses, including depression, psychosis, suicidal ideation, schizophrenia, across the lifespan (Doane et al. 2013; Heim et al. 2019; Jarcho et al. 2013; Miller et al. 2011; Taylor 2010; Vrshek-Schallhorn et al. 2013).

Additionally, recent evidence has also suggested that alterations in stress physiological systems (e.g. neuroendocrine, inflammatory, molecular, and structural pathways) due to developmental stress exposure may also alter sensitization to future stressors. The stress sensitization hypothesis (Hammen et al. 2000) proposes that the risk for adult mental illness following stressful life events is higher among individuals with a history of developmental trauma than among individuals without a history of developmental trauma. Recent findings have extended this hypothesis to suggest that greater early life trauma sensitizes or potentiates future reactions to stress and consequently, increases psychiatric disease risk (Heim et al. 2019; McLaughlin et al. 2010; Shapero et al. 2014). While an increasing number of studies are noting the long-term impacts of prenatal stress on infant, child, and early/mid adolescent psychological status (Ilg et al. 2019; Koss & Gunnar 2018; Ping et al. 2020; Ziljmans et al. 2015), few studies have examined the mental health impacts of prenatal stress into late adolescence and adulthood and also evaluated the potential stress sensitization effects of prenatal stress (Koss & Gunnar 2018; Ping et al. 2020; Ziljmans et al. 2015).

This emerging evidence suggests that some proportion of the mental health burden of current populations could reflect the lingering biological imprint of past traumatic experience during gestation (Barker 1999; Krontira et al. 2020; Kuzawa & Sweet 2009). South Africa’s recent history and the country’s tumultuous transition into democracy raise the question of whether the traumas of *Apartheid* may continue to have lasting effects, influencing risk for psychiatric morbidity across generations (Kim 2020). It furthermore raises the question of whether some of these effects might be reversible, opening opportunities for public health interventions to reduce the societal burden of psychiatric morbidity (Heim et al. 2019). However, little work in South Africa has explored this hypothesis and its contribution to mental health. Furthermore, as the most current research is conducted in high-income, Western contexts, it remains unclear if these effects are seen in non-Western, low- and middle-income contexts like South Africa, where adverse psychosocial and environmental conditions are more prevalent and where the burden of mental illness is substantially greater (Patel 2007; Vigo et al. 2016).

This study examines the long-term impacts of prenatal stress and trauma from South African *Apartheid* on psychiatric morbidity during late adolescence/early adulthood and the possible reversibility of the long-term effects of prenatal stress in a longitudinal birth cohort study. Adolescence is an important period in the development of future mental illness as most psychiatric conditions emerge during this stage (Patel 2007; Breslau et al. 2017) and because adolescent psychiatric disease is a major risk factor for future adult mental illness (Pine et al. 1993; Sawyer et al. 2012). The recent history of *Apartheid*, high rates of mental illness, and low rates of healthcare access emphasize the need to elucidate possible mechanisms underlying the intergenerational mental health effects of prenatal trauma in order to improve public mental health in South Africa.

## METHODS

### Study setting and participants

Data come from a longitudinal birth cohort in South Africa called Birth to Twenty Plus (Bt20++), which currently spans three generations of families in the greater Johannesburg-Soweto area. The Birth to Twenty (Bt20++) study is both the largest and longest running longitudinal birth cohort study of child health and development in Africa. Bt20++ emerged as a collaboration between the University of the Witwatersrand in Johannesburg and the South African Medical Research Council with the aim to assess the impacts of rapid urbanization towards the end of *Apartheid* on the growth, health, well-being, and educational progress of children. Soweto is the largest township outside of the city of Johannesburg and a site of immense cultural and historical significance in the struggle against *Apartheid*. Racial and political violence, government divestment, and widespread protest were common in both cities, particularly at the legislative end of *Apartheid*, which is when pregnant women were first recruited into the study.

All singleton live births delivered in public sector hospitals between April 23 to June 8, 1990 and who were residents in the metropolitan Johannesburg-Soweto area six months after delivery were enrolled in Bt20+ (Richter et al. 2007). In late 1989, BT20+ began interviewing pregnant women in public antenatal clinics to identify potential participants whose births would fall within the enrollment period. Enrolled Bt20+ neonates were cross-checked with all government birth notifications during the 7-week time period. The study area covered approximately 78 square miles at the time and included close to 3.5 million people, with about 400,000 informal housing units.

A total of 3273 singleton children were enrolled in Bt20+ and 72% of the initial cohort continued to participate in the study after 17 years (Norris et al. 2007). The cohort is roughly representative of the demographic parameters of the metropolitan Johannesburg-Soweto region. Currently, the cohort underrepresents White children due to cohort enrollment taking place in public health facilities; White families were more likely to utilize private practitioners and facilities Although BT20+ children were all *in utero* during the *Apartheid* regime, they became among the first generation born into a democratic South Africa, and colloquially known as “Mandela’s Children” because they were born shortly after Nelson Mandela’s release from prison on February 11, 1990. Between 1990 and 2007, the scope of this analysis, BT20+ families participated in 18 waves of data collection, and follow-up studies continue. All participants provided assent and their parents provided written, informed consent. Ethical approval was obtained from the University of Witwatersrand Committee for Research on Human Subjects.

Before the full sample was achieved, 1594 women were interviewed during their third trimester about their pregnancy, social experience, and household conditions. Antenatal interviews were conducted by seven trained, multilingual, interviewers. Where translation of measures was required, consensual agreement on the phrasing of questions was reached. Continuous translation and back-translation were used in order to ensure that the meaning/s attained in the destination language mirrored those intended in the original one. The majority of interviews were conducted in antenatal services, with a quarter conducted at home. Zulu, Sotho and English were the most common languages used. Adolescents were interviewed in a research facility at the Chris Hani-Baragwanath Hospital in Soweto and at home.

### Prenatal stress and social support

Prenatal stress exposure (G1) during the third trimester of pregnancy was collected using a 16-item scale (α = 0.64), adapted from Bluen et al. (1988) capturing information on exposure to various forms of family and community stresses, including police violence, injury, and incarceration. Yes/no responses indicated the presence or absence of each stressor during the previous 6 months. After exploratory analyses, two questions were dropped: one due to a low response rate (have you experienced any problems with your other children?) and the other due to contradictory concepts being assessed at once (domestic and familial violence vs. partner separation). Of the initial 1594 women, 1051 women (65.9%) completed all of the remaining prenatal stress questions.

Social support was measured using a series of four, yes/no questions to identify the absence or presence of instrumental and emotional support, including: people available to help, a confidante, being able to speak to her partner, belonging to a community organisation/church. All questions were summed to create a total score.

### Demographic, Health, and Socioeconomic Variables

During the antenatal visit in 1990, expectant women were asked about their current housing situation (e.g. number of rooms, number of inhabitants), population group (i.e. “race”), marital status, age, and obstetric history. Gravidity was calculated from information on obstetric history and operationalized into a categorical yes/no variable (0 = no past pregnancy, 1 = past history of pregnancy). They were also asked whether they used tobacco or had been drinking alcohol during their pregnancy. At delivery, a series of questions about the neonate were administered to gather information about gender, birth order, birthweight, and gestational age. Birthweight (g) and gestational age (weeks) were obtained from the child’s Road to Health Card, a patient-held child medical record provided to all new mothers in South Africa.

Household socio-economic status (SES) was assessed using an asset index which scored each participant according to the number of household physical assets that they possessed out of a possible 6 (e.g. television, refrigerator, washing machine, radio, telephone, home ownership, car). Household SES was measured again in 2006 using an updated list of assets (e.g. television, car, washing machine, refrigerator, phone, radio, microwave, cell phone, DVD, MNET, DSTV, computer, and internet). The asset index was designed based on standard measures used by the Demographic and Health Surveys (https://dhsprogram.com/), based on the work of Filmer and Pritchett (1999). To capture the overall SES environment during prenatal development and 2007, when the outcome measure was administered, an aggregate SES variable was constructed by summing standardized assets scores from both years of data collection.

### Household Stressful Life Events

The occurrence of stressful life events in the household were assessed across two timescales, six months and twelve months, and reported by the caregiver of the index child based on yes/no responses. Thirteen life events were assessed and summed together to create a composite measure of household stress that occurred during the past six months, which included death of a sibling, parent, and other family member (three separate questions), divorce, index child changing schools, a serious illness or hospitalization experienced by the index child, caregiver, and family member (three separate questions), marital separation, increase in arguments with partner, caregiver separation from family for two weeks or more, a child leaving home, and unemployment. Additionally, eight life events that occurred in the last year were summed to create a separate composite measure, which assessed whether any member of the household experienced robbery, harassment, threats, sexual molestation, physical violence, and murder.

### General Health Questionnaire (GHQ-28)

The General Health Questionnaire (GHQ-28) is a psychological screener that provides a measure of psychiatric morbidity based on four 7-item scales: somatic symptoms, anxiety and insomnia, social dysfunction, and severe depression (Goldberg & Hillier 1979). It assesses changes in mood, feelings, and behaviours during the past four weeks. The respondent evaluates their occurrence on a 4-point Likert scale, “less than usual,” “no more than usual,” “rather more than usual,” and “much more than usual.” Seven questions are reverse scored and transformed before all responses summed. Bt20+ index children and their caregivers completed the GHQ during a follow-up wave of data collection when the index children were 17-18 years old.

### Statistical analyses

All analyses were conducted using version 15.1 of Stata (Stata Corporation, College Station, TX). All variables were examined for normal distribution and outliers. Bivariate analyses were conducted between prenatal stress, psychiatric morbidity at age 18, and covariates. Covariates were included based on a priori knowledge of social, biological, and obstetric risk factors that may potentially confound the relationship between prenatal stress and later-life psychiatric morbidity (de Mola et al. 2014; Entringer et al. 2009; O’Donnell et al. 2013). The following variables were considered for inclusion as confounding factors: maternal age, maternal education, marital status, gravidity, tobacco use, and alcohol consumption during pregnancy, household density (ratio of the number of inhabitants to the number of rooms available for sleeping), social support, and child gender. (*p* > 0.10). Fetal growth rate and gestational age were also included as key covariates to examine the role of fetal growth restriction and preterm delivery as possible pathways by which prenatal stress affects late adolescent psychiatric morbidity. A proxy measure of fetal growth was created by regressing birthweight on gestational age to create standardized residuals.

Bivariate analyses were conducted to identify potential confounding variables for inclusion in the final analytic models. With the exception of known confounding factors for the relationship between prenatal stress and later life psychiatric morbidity, only those that were statistically significant at the 0.1 level were included in the final models. These variables included infant gender, aggregate SES, gravidity, perceived social support during pregnancy, maternal education during pregnancy, household density, alcohol consumption during pregnancy, tobacco use during pregnancy, fetal growth rate, gestational age, maternal age, recent stress, past stress, and maternal GHQ in 2007. Multiple ordinary least squares (OLS) regressions were conducted to examine the impact of prenatal stress on late adolescent psychiatric morbidity.

The final analytical sample included 304 mothers and adolescents with requisite data (Table 1). Participants included in the analytical sample were similar to those excluded (n = 747) with respect to psychiatric morbidity at 17-18 years, prenatal stress, child gender, gravidity, alcohol consumption, tobacco use, social support, household density, SES in 2008, fetal growth rate, recent stress, past stress, and maternal psychiatric morbidity (*p* > 0.05). Birthweight, gestational age, SES in 1990, maternal education in 1990, and the aggregate SES measure were significantly different from those excluded from the sample (*p* < 0.05). Children in the analytical sample exhibited higher birthweights and slightly longer gestations. Additionally, participants in the analytical sample reported lower household SES in 1990, lower aggregate SES levels, but more educated mothers in 1990. Thus, the analytic sample is somewhat disadvantaged compared to excluded group.

**Table 1.**
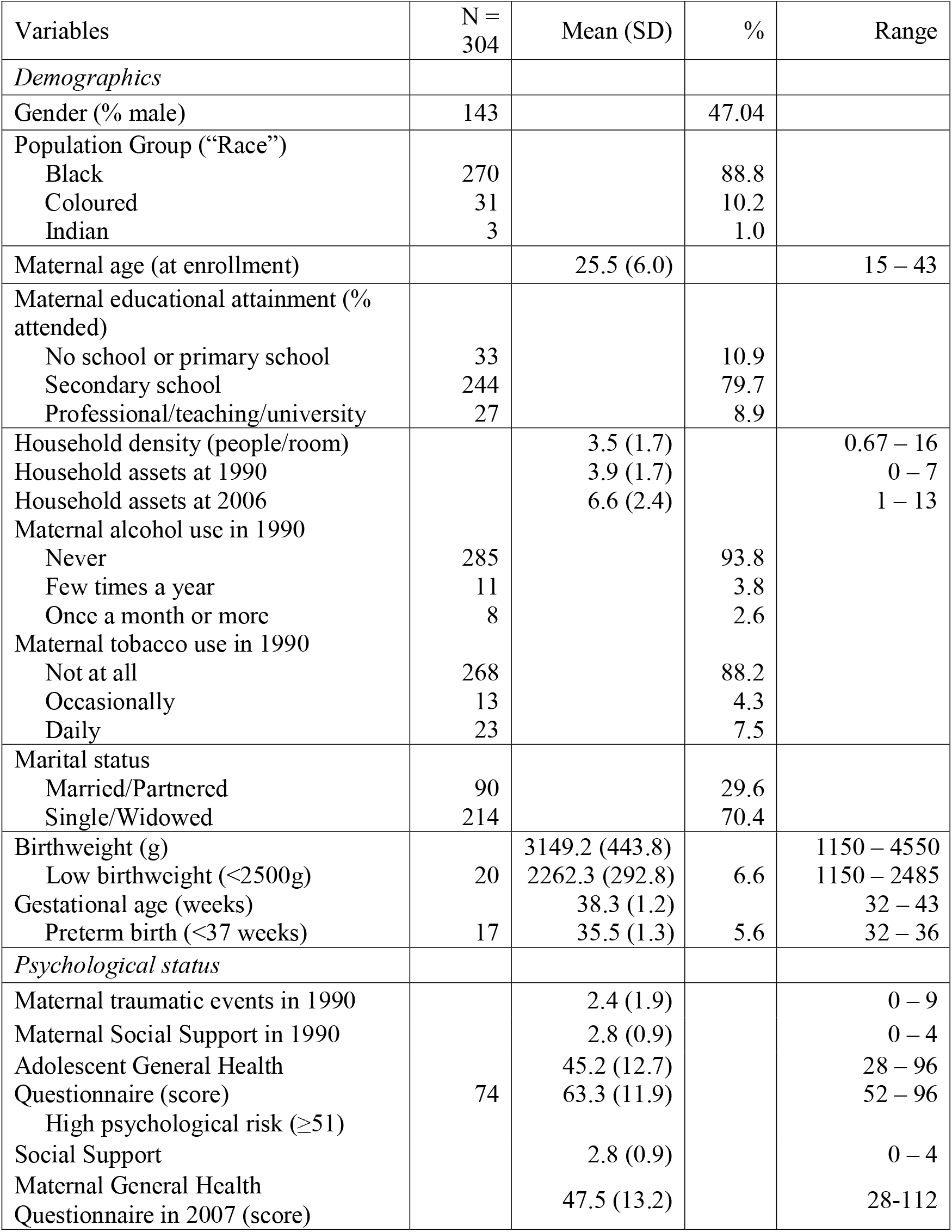
Demographic characteristics, prenatal conditions, and psychiatric morbidity

## RESULTS

In our sample of 304 mothers and adolescent pairs, the average number of traumatic events experienced within the past six months of the interview was 2.4/15 total events, while the average number of social support resources was 2.8/4 (Table 1). About 24% of the sample, or 74 adolescents, surpassed the cutoff (51/112) for high psychological morbidity based on their GHQ scores. Table 2 presents the results of the OLS regression analyses of fetal, maternal, behavioral, and environmental factors that predict psychiatric morbidity during late adolescence. The unadjusted model (Model l) predicting GHQ scores on prenatal stress was positive although not significant (β = 0.53, *p* = 0.169). The effect of prenatal stress did not substantively change after adjusting for gender or maternal age (Model 2). We tested for an interaction between prenatal stress and gender, but we found no evidence for gender differences. The relationship between prenatal stress and psychiatric morbidity strengthened and approached significance (β = 0.69, *p* = 0.066) after including gestational variables, specifically gravidity, gestational age, and fetal growth (birthweight adjusted for gestational age) (Model 3). Greater fetal growth rates significantly predicted greater adolescent GHQ scores (β = 0.035, *p* = 0.037). Adjusting for maternal pregnancy exposures (Model 4), stress measures and maternal GHQ (Model 5), socioeconomic markers (Model 6), and prenatal social support (Model 7) did not appreciably modify the relationship between prenatal stress and adolescent GHQ, but the effect of prenatal stress falls out of significance in Model 6.

**Table 2.** Multiple regression models of prenatal stress predicting adolescent psychiatric morbidity with covariates

|  | Model 1 | Model 2 | Model 3 | Model 4 | Model 5 | Model 6 | Model 7 |
| --- | --- | --- | --- | --- | --- | --- | --- |
| Prenatal stress (count) | 0.5 ± 0.4 | 0.6 ± 0.4 | 0.7 ± 0.4<sup>†</sup> | 0.7 ± 0.4<sup>†</sup> | 0.7 ± 0.4<sup>†</sup> | 0.6 ± 0.4 | 0.6 ± 0.4 |
| Gender (male) |  | -5.9 ± 1.4*** | -6.0 ± 1.4*** | -6.0 ± 1.4*** | -5.9 ± 1.5*** | -5.9 ± 1.5*** | -5.8 ± 1.5*** |
| Maternal age (year) |  | -0.1 ± 0.1 | -0.01 ± 0.1 | -0.02 ± 0.1 | 0.03 ± 0.2 | 0.02 ± 0.2 | 0.04 ± 0.2 |
| Gravidity (yes/no) |  |  | -3.1 ± 1.8<sup>†</sup> | -3.1 ± 1.8<sup>†</sup> | -3.3 ± 1.9<sup>†</sup> | -3.9 ± 1.9* | -3.7 ± 1.9<sup>†</sup> |
| Fetal growth<sup>a</sup> |  |  | 0.004 ± 0.002* | 0.003 ± 0.002* | 0.003 ± 0.002<sup>†</sup> | 0.003 ± 0.002<sup>†</sup> | 0.003 ± 0.002<sup>†</sup> |
| Gestational age (week) |  |  | -0.1 ± 0.6 | -0.1 ± 0.6 | -0.1 ± 0.6 | -0.2 ± 0.6 | -0.2 ± 0.6 |
| Alcohol (frequency) |  |  |  | 0.01 ± 0.9 | 0.04 ± 0.9 | 0.002 ± 0.9 | -0.09 ± 0.9 |
| Tobacco (frequency) |  |  |  | 0.7 ± 1.3 | 0.7 ± 1.3 | 0.3 ± 1.4 | 0.1 ± 1.4 |
| Crowding (ratio) |  |  |  |  | -0.3 ± 0.4 | -0.4 ± 0.4 | -0.4 ± 0.4 |
| Recent Stress (count) |  |  |  |  | 1.4 ± 0.8<sup>†</sup> | 1.5 ± 0.8<sup>†</sup> | 1.5 ± 0.8<sup>†</sup> |
| Past Stress (count) |  |  |  |  | 1.0 ± 1.2 | 1.1 ± 1.2 | 1.0 ± 1.2 |
| Maternal GHQ (score) |  |  |  |  | -0.02 ± 0.06 | -0.03 ± 0.06 | -0.04 ± 0.06 |
| Assets (count) |  |  |  |  |  | -0.3 ± 0.5 | -0.2 ± 0.5 |
| Maternal education (count) |  |  |  |  |  | -0.9 ± 0.9 | -0.8 ± 0.9 |
| Social support (count) |  |  |  |  |  |  | -0.9 ± 0.9 |
| Intercept | 43.9 ± 1.2*** | 50.2 ± 3.3*** | 54.0 ± 22.4* | 52.1 ± 22.8* | 53.8 ± 23.3* | 59.4 ± 24.2* | 62.9 ± 24.4* |
| Model R<sup>2</sup> | 0.0063 | 0.0641 | 0.0858 | 0.0866 | 0.0986 | 0.1037 | 0.1071 |

<sup>†</sup> $p < 0.1$ \* $p < 0.05$ \*\* $p < 0.01$ \*\*\* $p < 0.001$ <sup>a</sup> Birthweight adjusted for gestational age2

|  | Model 8 | Model 9 |
| --- | --- | --- |
| Prenatal stress (count) | 4.8 ± 1.6** | 0.1 ± 0.5 |
| Prenatal stress x Maternal age | -0.2 ± 0.1** |  |
| Prenatal stress x Past Stress |  | 1.2 ± 0.6* |
| Gender (male) | -5.7 ± 1.4*** | -5.7 ± 1.5*** |
| Maternal age (year) | 0.4 ± 0.2* | 0.7 ± 0.2 |
| Gravidity (yes/no) | -3.6 ± 1.9<sup>†</sup> | -3.8 ± 1.9<sup>†</sup> |
| Fetal growth<sup>a</sup> | 0.003 ± 0.002* | 0.003 ± 0.002<sup>†</sup> |
| Gestational age (week) | -0.1 ± 0.6 | -0.2 ± 0.6 |
| Alcohol (frequency) | -0.004 ± 0.9 | -3.8 ± 1.9 |
| Tobacco (frequency) | 0.1 ± 1.4 | -0.1 ± 1.4 |
| Crowding<sup>b</sup> (ratio) | -0.3 ± 0.4 | -0.3 ± 0.4 |
| Recent Stress (count) | 1.5 ± 0.8<sup>†</sup> | 1.5 ± 0.8<sup>†</sup> |
| Past Stress (count) | 0.9 ± 1.2 | -1.6 ± 1.7 |
| Maternal GHQ (score) | -0.03 ± 0.1 | -0.04 ± 0.06 |
| Assets (count) | -0.3 ± 0.5 | -0.1 ± 0.5 |
| Maternal education (count) | -0.7 ± 0.9 | -0.7 ± 0.9 |
| Social support (count) | -1.1 ± 0.9 | -0.9 ± 0.9 |
| Intercept | 50.9 ± 24.6* | 61.7 ± 24.3* |
| Model R<sup>2</sup> | 0.1288 | 0.1210 |

<sup>†</sup> $p < 0.1$ \* $p < 0.05$ \*\* $p < 0.01$ \*\*\* $p < 0.001$ <sup>a</sup> Birthweight adjusted for gestational age <sup>b</sup> Number of household inhabitants/rooms used for sleeping

### Testing moderators of prenatal stress-later life psychiatric morbidity pathway

We next assessed factors that could moderate the relationship between PNS and future psychiatric morbidity in the index generation: maternal age, recent/past stress, and social support. We examined the effect of an interaction between gender and prenatal stress on psychiatric morbidity, and results showed a non-significant interaction between prenatal stress and male (β = -0.46, *F*[1, 287] = 0.35, *p* = 0.556). Model 8 reports a negative and significant interaction between maternal age and prenatal stress severity (β = -0.17, *F*[1, 287] = 7.14, *p* = 0.008), showing that the adverse psychiatric effects of prenatal stress are stronger in children with younger mothers (Figure 2).

**Figure 1.**
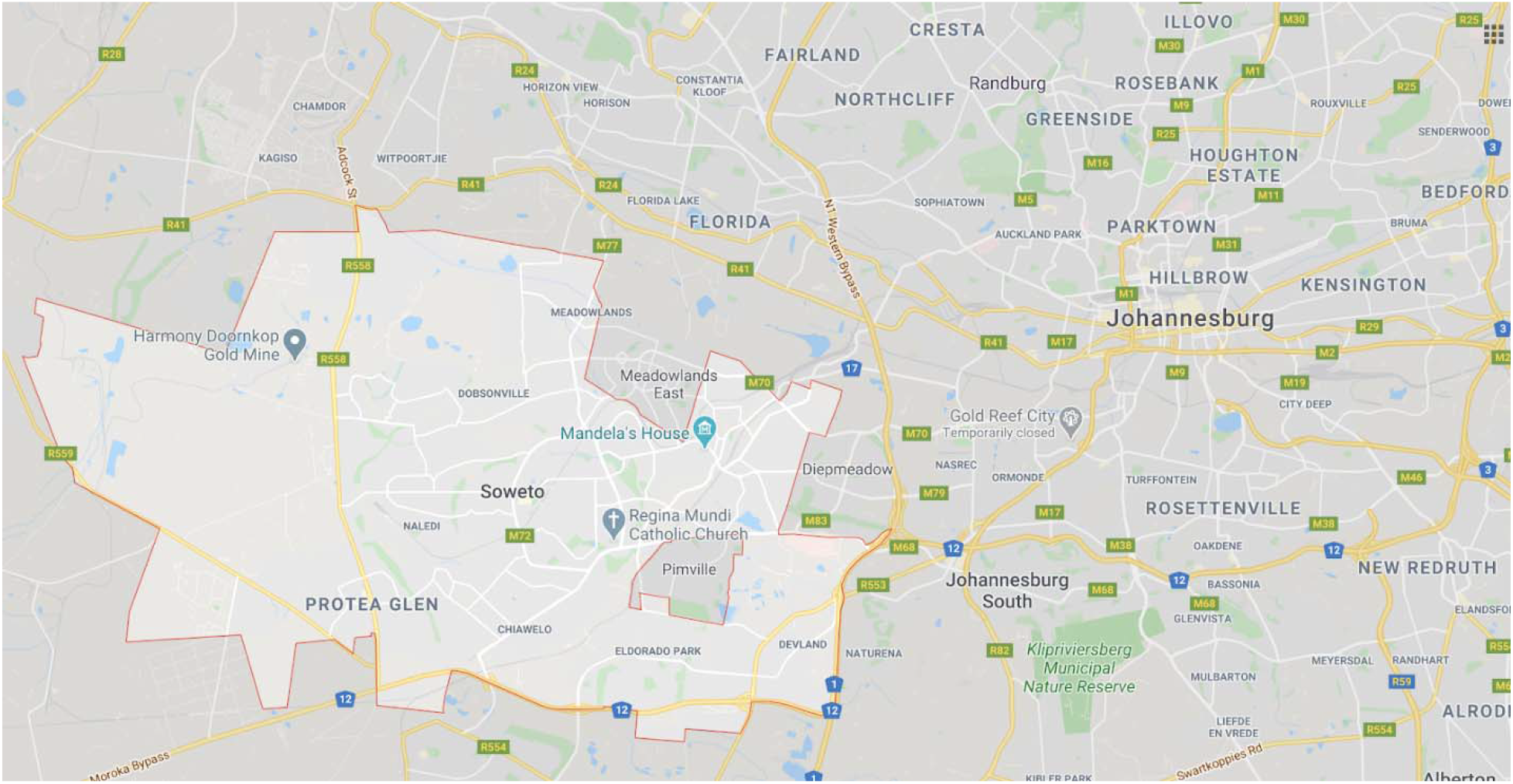
Map of Johannesburg and Soweto

**Figure 2.**
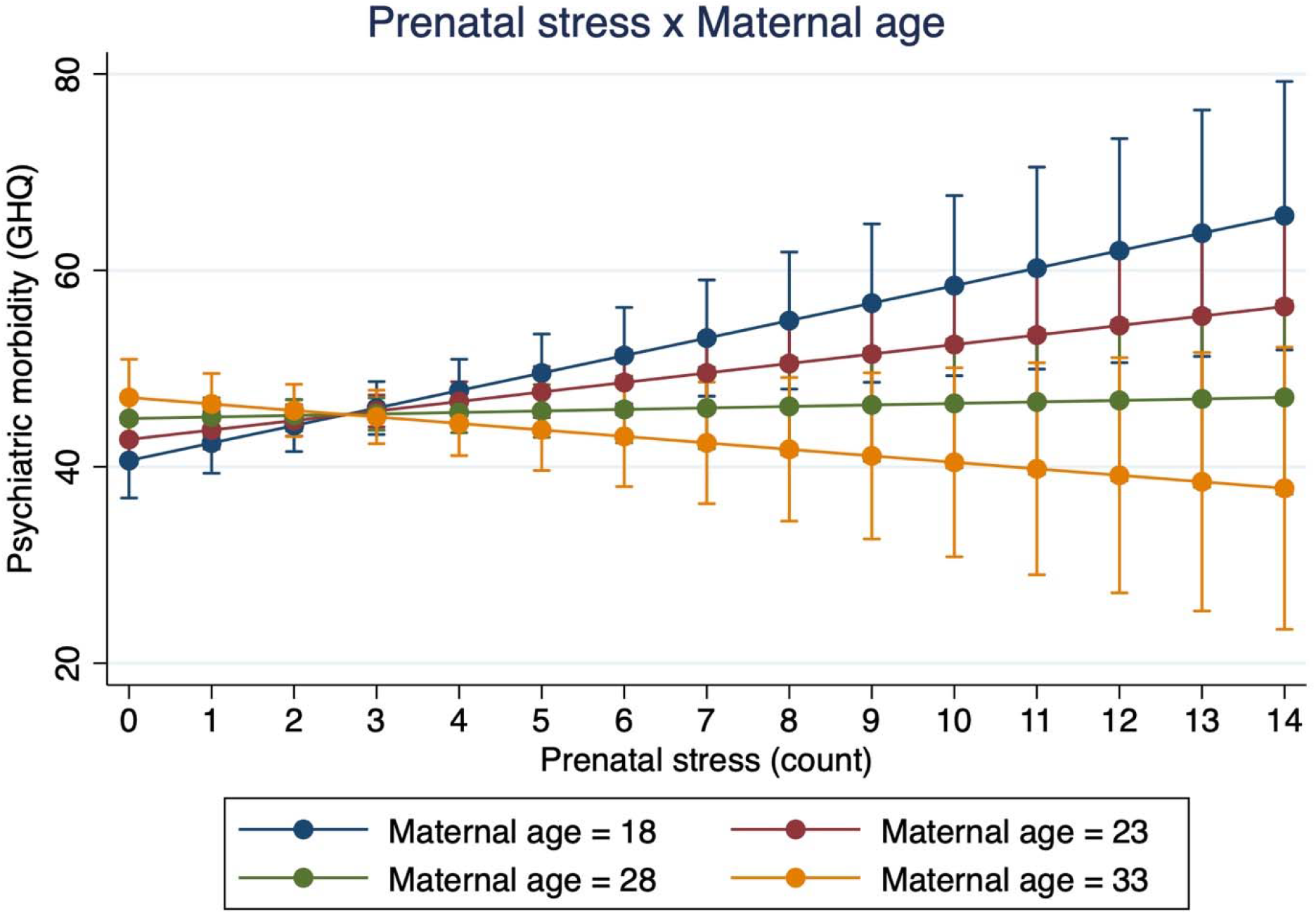
*Interaction effect between prenatal stress and maternal age predicting psychiatric risk*. *Note*. This figure demonstrates the interaction effect between prenatal stress and maternal age predicting adolescent psychiatric risk at Year 17. The effect of prenatal stress on late adolescent psychiatric risk is much stronger among younger mothers (β = -0.17, *F*[1, 287] = 7.14, *p* = 0.008). There were significant main effects for prenatal stress and maternal age.

To assess the stress-sensitization hypothesis, we evaluated the interaction between prenatal stress and both measures of household stress from the past six and twelve months. Our results reported a non-significant interaction between prenatal stress and recent household stress from the past six months (β = - 0.027, *F*[1, 287] = 0.001, *p* = 0.951) and a significant interaction between prenatal stress and past household stress from the past year (Model 9: β = 1.19, *F*[1, 287] = 4.52, *p* = 0.0343) Figure 3). The effect of prenatal stress on late adolescent and early adult psychiatric morbidity was stronger in index children living in households with greater stress and trauma exposure. Finally, to explore the possible effects of positive maternal social environments during pregnancy in ameliorating the long-term mental health impacts of prenatal stress, we evaluated the effect of an interaction between social support and prenatal stress on psychiatric morbidity. Results reported a non-significant interaction between social support and prenatal stress (β = -0.092, *F*[1, 287] = 0.04, *p* = 0.836).

**Figure 3.**
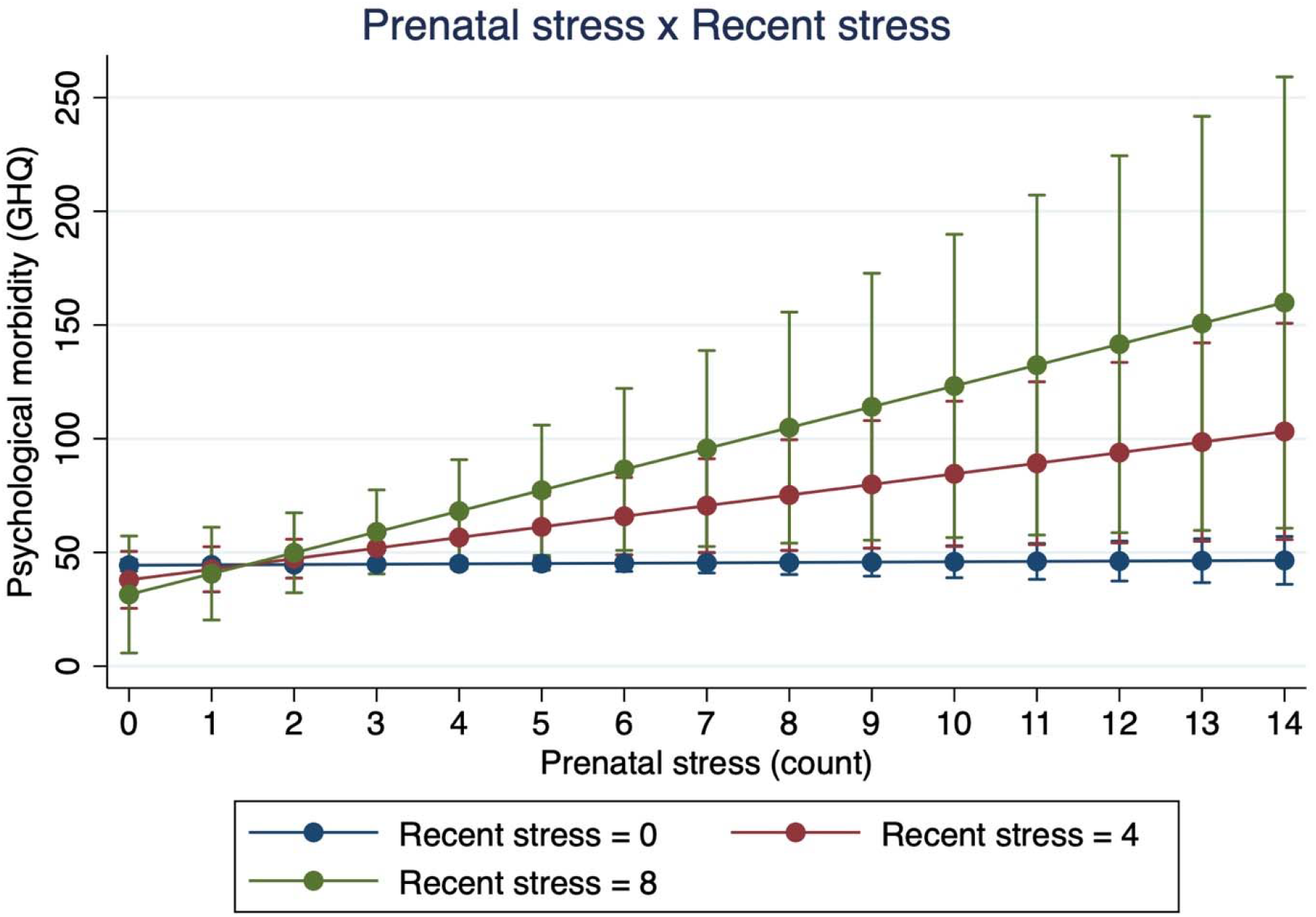
*Interaction effect between prenatal stress and household stress and trauma in the past year predicting psychiatric risk. Note*. This figure demonstrates the interaction effect between prenatal stress and recent stress predicting adolescent psychiatric risk at Year 17. The effect of prenatal stress on late adolescent psychiatric risk is stronger as the degree of recent stress increases (β = 1.19, *F*[1, 287] = 4.52, *p* = 0.0343). There were no significant main effects for prenatal stress and recent stress.

## DISCUSSION

In this longitudinal study of the intergenerational mental health impacts of prenatal stress in South Africa, we find that pregnant women who reported greater trauma exposure during *Apartheid* had children who exhibited greater psychiatric morbidity during late adolescence, 17-18 years after the timing of their fetal stress exposure. This relationship, however, was only significant after interacting greater prenatal stress with younger maternal age (*p* = 0.008) and greater household stress and trauma in the past year (*p* = 0.0343) and remained statistically significant after controlling for key demographic, social, and biological characteristics. In sum, these data shed light on the potential fetal origins of late adolescent mental health and the intergenerational effects of trauma from *Apartheid* in our birth cohort sample of South African mothers and children in Soweto-Johannesburg. This study is among the first to prospectively assess the long-term psychiatric impacts of prenatal stress into early adulthood in a low- and middle-income country.

The finding that greater prenatal stress predicts future psychiatric morbidity is consistent with the growing literature on the fetal origins of later life psychopathology (Abbott et al. 2018; Bosch et al. 2012; Davis et al. 2020; O’Donnell et al. 2013; Ping et al. 2020). Retrospective and prospective studies of prenatal stress show that greater maternal social adversity during pregnancy predicts elevated risks for developing psychopathologies like depression, psychosis, and schizophrenia in the future (Lipner et al. 2019; McQuaid et al. 2019; Van den Bergh et al. 2008). The long-term, intergenerational effects of prenatal stress, however, were only significant when the moderating effects of maternal age and past traumatic life events on prenatal stress were independently assessed.

Our findings on the interaction between prenatal stress and younger maternal age corroborate past research highlights the numerous socioeconomic, gendered, and cultural adversities faced by younger pregnant mothers and their families in Soweto and other communities in South Africa (Makola 2011; McLeod 1999; Richter et al. 2006; Willan 2013). Young motherhood in Soweto is understood to be a highly stigmatized and morally compromised status in Soweto and a period of greater vulnerability to certain forms of violence in Soweto and other communities in South Africa (Panday et al. 2009). Past research in Bt20+, Soweto, and elsewhere shows that young mothers frequently receive disappointment and negative attitudes from their parents, ridicule and shame from nurses, and stigma from community members (Makola 2011; McLeod 1999; Richter et al. 2006; Willan 2013) and that infants of young teenage mothers were lighter at birth relative to neonates from older mothers (Cameron et al. 1996; Lundeen et al. 2016; Rothberg et al. 1991). Our data also showed that both younger age was associated with lower birthweight. The long-term developmental effects of lower birthweight are well-documented – lower birthweight is associated with increased risk for a wide range of adolescent and adult mental illness risk across the lifecourse (Abel et al. 2010; Barker 2004; Lærum et al. 2019; Orri et al. 2019).

Results also show that fetal growth, but not gestational age, is an important predictor of later life psychiatric morbidity in this sample. Fetal growth was positively and significantly related to late adolescent/early adulthood psychiatric morbidity. Also, our finding that fetal growth weakens the coefficient on prenatal stress after inclusion into the model provides preliminary evidence that fetal growth may be involved in the relationship between prenatal stress and adolescent psychiatric morbidity. The direct effect of fetal growth rate on psychiatric morbidity at 18, however, conflicts with past literature on the fetal origins hypothesis that suggests that adverse intrauterine exposures and slower fetal growth, potentially due to stress-induced alterations in gestational neuroendocrine activity, can durably impact health, development, and disease risk in the next generation (Kuzawa 2008; O’Donell & Meaney 2017; Monk et al. 2019). Past studies have shown sex differences in fetal growth rates and preterm birth with boys typically showing higher incidence of adverse birth outcomes (Di Renzo et al. 2007), yet there were no significant sex differences in fetal growth rates, gestational age, nor birthweights in this sample. Further research in Bt20+ should examine the potential sex differences in fetal growth rates due to prenatal stress.

Our data show that recent stress from the past 6 months was a positive and marginally significant predictor of psychiatric morbidity at 17-18, while the effect of traumatic life experiences from the past 12 months was only significant when interacted with prenatal stress. The significant interaction between prenatal stress and past year traumatic experiences, showing that the direct relationship between prenatal stress and psychiatric morbidity is stronger in children with greater household traumatic events, provide supporting evidence for the stress sensitization hypothesis (Hammen et al. 2000; McLaughlin et al. 2010; Van den Bergh et al. 2008). While the data show that prenatal stress itself may not have sensitizing effects on later life psychiatric morbidity, it is only after interacting with elevated levels of past stress that the long-term sensitizing effects of prenatal stress on later life psychiatric morbidity become apparent. The stress sensitizing effect of prenatal stress may only significantly interact with past stress (from the past 12 months) trauma rather than with recent stress (from the past six months) because of the nature of the experiences queried by each measure. Reported by the mother during the interview, our composite measure of past stress consisted of eight questions assessing a collection of objective traumatic events that are more likely to have household-level impacts specifically on the index child compared to the events assessed in the recent stress variable. Also reported by the mother, our measure of recent stress may also query household experiences that may have marginal to no impact on the index child’s psychiatric morbidity. These experiences include unemployment, divorce, or death of a relative. Nevertheless, our data show interesting stress sensitization effects due to the interaction between prenatal stress and household stress exposure from the past year.

Given these data and existing findings on the fetal origins of late adolescent/early adult psychopathology, there are two possible developmental mechanisms that may facilitate the lasting impacts of prenatal stress on future psychiatric morbidity. First, increased severity of prenatal stress may cause durable increases in psychological and physiological stress reactivity (e.g. HPA axis, the immune system, and brain function) into adulthood, which may make individuals respond worse to future stressors, and in turn increase one’s risk of developing a psychopathology (Hammen et al. 2000; Heim et al. 2019; Kendler et al. 2004; McLaughlin et al. 2010). In one study, researchers found that women who experienced greater maternal anxiety during pregnancy were more likely to have children with flattened diurnal cortisol slopes, which predicted depression in female adolescents (Van den Bergh et al. 2008) The growing literature on the long-term health effects of prenatal stress is consistent with the larger body of scholarship on the effects of postnatal early life stress, which are known to have similar lasting effects on neuroendocrine, inflammatory, and molecular mechanisms across development, extending into adulthood (Gustafsson et al. 2010; Heim & Binder 2012; Taylor et al. 2010).

Second, greater histories of prenatal stress may increase the severity of behavioral and psychiatric conditions that increase emotional and biological sensitization to future stressors and adverse events, such as depression, anxiety, and other mood disorders. Additionally, the major symptoms of depression, such as persistent feelings of victimization, learned hopelessness and helplessness, and negative appraisal (Folkman & Lazarus 1986; Peterson & Seligman 1983) may have elevated individual sensitivity to and appraisal of recent stressful events (Medrano & Hatch 2005; Peterson & Seligman 1983) and neuroendocrine sensitization (Stroud et al. 2011) to future stressors. Thus, greater sensitivity to and appraisal of past stress may have emerged as a function of the long-term depressive and psychological effects of prenatal stress. Future longitudinal research is needed to determine the underlying stress physiological mechanisms by which prenatal stress influences future stress sensitivity and psychopathological morbidity.

Additional stress physiological mechanisms and external social processes may also explain the late adolescent psychiatric impacts of prenatal stress in our sample. Growing research report that higher levels maternal prenatal stress corresponds with greater pro-inflammatory cytokines levels and other inflammatory markers across adulthood (Bilbo & Schwarz 2009; Entringer et al. 2008; Plant et al. 2016; Slopen et al. 2015). Scientists have also found increasing evidence that greater maternal prenatal stress and social experience predicts epigenetic changes at genetic loci affecting neuroendocrine systems, neurotransmission, and neurogenesis in their children (Barker et al. 2018; Glover et al. 2018; Ostlund et al. 2016; Provenzi et al. 2020). While we cannot completely rule out the potential role of certain stress physiological mechanisms in driving the prenatal stress-late adolescence relationship without directly assessing the system (e.g. salivary cortisol, cytokines, etc.), multiple stress-sensitive biological pathways likely contribute to elevations in psychiatric risk at the same time. Finally, prenatal stress may be indicative of larger and longer-term socioeconomic patterns of mothers and children in Soweto. Substantial evidence illustrates the durable impacts of poverty and material deprivation during pregnancy. Chronically low socioeconomic status between gestation and adulthood predicted flatter diurnal cortisol rhythms in a large sample of Filipino adults (DeSantis et al. 2015).

We also report that perceived social support during pregnancy did not buffer against the later-life effects of trauma exposure during pregnancy. Prenatal social support did not appear to have an independent protective effect on adolescent psychiatric morbidity, yet the presence of a partner during pregnancy did significant predict lower adolescent GHQ scores. The insignificant effect of prenatal social support on future psychiatric morbidity, either from a direct protective effect on adolescent mental health or buffering against the psychiatric impacts of prenatal stress, may be attributed to the limited strength of our social support measure, which primarily assessed interpersonal support and a single, dichotomous measure of group membership.

Psychologists have emphasized the importance of assessing the degree, frequency, and timing, and modes of social support, similar to the impacts of stress and trauma, when examining psychosocial and physiological impacts (Dunkel Schetter 2011; Orr 2004) Past studies report protective effects of prenatal social support on postnatal outcomes, including better infant birth outcomes (Feldman et al. 2000; Orr 2004) and lower child adiposity (Katzom et al. 2019). Prenatal social support, in the form of a presence of a partner during pregnancy, has also shown to buffer the adverse effects of prenatal social adversity to predict better cognitive and psychiatric outcomes in children (Spann et al. 2020). Finally, early evidence suggest that prenatal social support may buffer against the impacts of maternal stress and contribute to healthier cortisol regulation (Field et al. 2013; Luecken et al. 2013). Further research is necessary to identify both the protective and buffering effects of prenatal stress to ameliorate the long-term disease outcomes of intergenerational stress and trauma.

In many past published studies of the lifecourse impacts of adverse prenatal experience, the source of maternal social adversity stem from violent and oppressive conditions linked with political turmoil, war, famine, and numerous forms of social inequality (Yehuda et al. 2016; Kertes et al. 2016; Roseboom et al. 2006; Kim et al. 2020). Despite the formal end to these forms of political violence, the developmental and health consequences of trauma exposures among pregnant women can durably extend across the lifecourse of the future child and may even impact the subsequent generation (Barbarin & Richter 2013; Kuzawa & Sweet 2009). These developmental pathways and health consequences of embodied trauma may represent mechanisms that drive mental health inequalities that disproportionately impact marginalized populations with high levels of stress and trauma exposure. Inasmuch as scientists aim to reverse the past impacts of embodied trauma and social oppression, the ongoing legacies of these violent histories and historical traumas, such as *Apartheid*, must be recognized and addressed to prevent future mental health inequities from emerging.

## LIMITATIONS

Unfortunately, the timeframe of the queried stress exposure in the prenatal stress measurement does not specify the exact moment of exposure as the participants were asked to report stressors that occurred in the past six months. Women completed the antenatal stress questionnaire during their third trimester, meaning that the reported stressor could have occurred anytime between the first and third trimester. Future research will benefit from knowing the exact periods of prenatal stress and cortisol exposure to understand the potential developmental and physiological effects on the child.

## CONCLUSION

In this analysis of the intergenerational mental health impacts of prenatal stress in a large birth cohort in Soweto, South Africa, our data show that pregnant women who reported greater trauma exposure during *Apartheid* had children who exhibited greater psychiatric morbidity during late adolescence and early adulthood, 17-18 years after the timing of their fetal stress exposure. This relationship, however, was only significant among children born to younger mothers and children exposed to household adversity in the past year. These findings suggest that greater prenatal stress may adversely affect adolescent mental health, have stress-sensitizing effects in children, and represent possible intergenerational effects of trauma experienced under *Apartheid* in this sample of South African mothers and children in Soweto-Johannesburg.

## Data Availability

Data are available upon request to the Developmental Pathways for Health Research Unit at the University of the Witwatersrand.

